# Increasing Lung Cancer Screening Participation Using an Informational Video Nudge: A Randomized Feasibility Trial

**DOI:** 10.64898/2026.08.28.26361654

**Authors:** Kris F. Wain, Nikki M. Carroll, Brian Hixon, Julie Steiner, Andrew J. Maclennan, Debra P. Ritzwoller

## Abstract

**Purpose:** Lung cancer screening (LCS) with low-dose computed tomography (LDCT) reduces lung cancer mortality, yet screening participation remains low. We evaluated whether a brief informational video nudge delivered immediately before a scheduled clinical encounter increased LCS ordering and baseline LCS completion.

**Patients and Methods:** We conducted a randomized feasibility trial within Kaiser Permanente Colorado from March through October 2025. LCS-eligible patients with an upcoming primary care or pulmonology appointment were assigned to intervention or usual care based on birth month. Intervention patients were split into two group, a group who received the LCS informational video nudge via text message within 24 hours of an eligible appointment; and second group who received the text plus a QR code video link during appointment rooming. Outcomes included LCS orders, baseline LCS-LDCT completion, and video engagement. Multivariable logistic regression was used to evaluate factors associated with LCS ordering.

**Results:** Among 1,093 patients, 549 were assigned to intervention and 544 to usual care. Intervention patients were more likely to receive an LCS order within 1 day of their appointment (22.6% vs 16.4%; *p*=.010) and any time during follow-up (32.6% vs 24.1%; *p*=.002). Baseline LCS-LDCT completion was 51% higher in the intervention group, although the difference was not statistically significant (8.6% vs 5.7%; *p*=.078). Among the intervention group, 93 individuals (17%) viewed the video, generating 114 total views, and viewers watched an average of 79% of the video. Most views (82.5%) occurred through text-message delivery rather than QR codes.

**Conclusion:** A brief, low-burden LCS informational video delivered immediately before a clinical encounter and integrated into existing workflows significantly increased LCS ordering and was associated with higher screening completion. Timely, scalable digital nudges may provide an effective strategy for improving LCS participation. Based on the observed effectiveness, feasibility, and efficiency of the intervention, KPCO incorporated the behavioral nudge into standard clinical care in February 2026.

## Introduction

Lung cancer screening (LCS) with low-dose computed tomography (LDCT) has been shown to reduce lung cancer mortality by 20% among high-risk individuals.^1^ Despite the noted benefits, participation in LCS programs remains low, with only 16% to 37% of eligible individuals up to date for screening.^2–6^ Healthcare information technology has proven effective in influencing patients’ decisions through the use of “nudges”.^7^ Informational nudges, such as brief videos emphasizing the benefits of LCS, may lead to improved screening particiaption.^8^

In contrast to other U.S. Preventive Services Task Force (USPSTF) recommended cancer screenings, LCS requires shared decision-making (SDM) between the patient and provider to discuss the benefits and risks before it can be ordered.^9^ However, many patients do not engage in meaningful screening discussions with their healthcare providers because of patient- and provider-level barriers, including limited time, competing clinical priorities, financial concerns, and variability in knowledge and attitudes toward screening.^10^ Well-designed nudge interventions can address both patient-level and provider-level barriers to cancer screening, improve the quality of SDM discussions, and ultimately increase participation.

The effectiveness or prior nudge interventions has often been context dependent and shaped by factors such as timing, relevance to the target behavior, integration into existing clinical workflows, and the credibility and visibility of the message.^11^ In our prior work, we found that pre-visit nudges delivered to LCS-eligible patients through targeted electronic and mailed outreach did not significantly increase LCS participation.^12^ Other interventions have found that LCS informational video improved patient knowledge around screening, but did not increase LCS participation.^13^ Interventions that have focused on selected populations, such as current smokers undergoing smoking cessation, have found significant increases in LCS following an informational video intervention.^14^ Collectively, these findings suggest that informational content alone may be insufficient and that the timing, delivery mechanism, and clinical context of the intervention are critical to its effectiveness.

Leveraging and extending upon our prior findings, the purpose of this feasibility study is to evaluate whether enhancing our prior pilot intervention by delivering a brief LCS informational video via text message within 24 hours of a scheduled clinical appointment and embedding the outreach within existing appointment-reminder workflows can improve rates of provider initiated LCS orders and patient completion of screening. This approach is designed to reach patients at a particularly actionable moment, immediately before a clinical encounter, when they are positioned to discuss LCS with their provider and act on screening recommendations. By combining timely outreach, concise educational content, and integration into routine clinical workflows, the intervention is designed to improve shared decision-making and increase LCS participation while minimizing additional burden on clinicians and health system resources. Beyond improving screening participation, this approach may also help health systems proactively address emerging quality measures related to LCS, including forthcoming HEDIS metrics.^15^ Because the intervention is intentionally designed to be low resource, scalable, and readily integrated into existing communication infrastructure, it has the potential for broad dissemination across healthcare settings and populations.

## Methods

### Study Setting and Participants

This study was conducted within Kaiser Permanente Colorado (KPCO), an integrated health maintenance organization serving more than 500,000 members. The study population was restricted to KPCO members receiving care within the Denver/Boulder service area. Approximately 10,000 KPCO members were eligible for lung cancer screening (LCS) during the study period, and historical completion of LCS with low-dose computed tomography (LDCT) among LCS-eligible members was approximately 37%. KPCO members routinely receive text-message reminders for scheduled primary care appointments, providing an established communication infrastructure for intervention delivery. In addition, KPCO has implemented a “care-gap” reminder for patients who met 2021 U.S. Preventive Services Task Force LCS eligibility criteria but had never completed screening. This care-gap infrastructure provided a defined population of LCS-eligible patients for targeted outreach.

The Kaiser Permanente Interregional Institutional Review Board determined that this quality improvement initiative did not meet the regulatory definition of research; therefore, informed consent was not required. This pragmatic quality-improvement feasibility trial was not prospectively registered ClinicalTrials.gov because it was conducted as a health-system quality-improvement initiative and was determined by the institutional review board not to meet the regulatory definition of research.

### Cohort identification and randomization

Between March 27, 2025 and October 31, 2025, daily EHR data was used to identify all patients with an active LCS care gap who had a primary care or pulmonology visit scheduled for that day or the next day. Patients were randomized by birth month: those with an odd birth month (e.g., January, March, etc.) were randomized to the Intervention Group, which was further divided into two delivery modalities: text-message only and text-message plus QR code based upon the patients bonded primary care clinic location. Patients with an even birth month (February, April, etc.) were randomized to Usual Care (Figure 1). Patients were followed from their first completed appointment to November 30, 2025, which was the end of the follow-up period. Patients in the intervention group received the nudge video before each eligible appointment. Outreach was discontinued once a patient either completed a baseline LCS-LDCT or indicated that they did not wish to participate in LCS. Individuals were excluded if they had a prior lung cancer diagnosis, were receiving hospice care, had opted out of text-message outreach, or did not have a qualifying primary care or pulmonology appointment.

**Figure 1.**
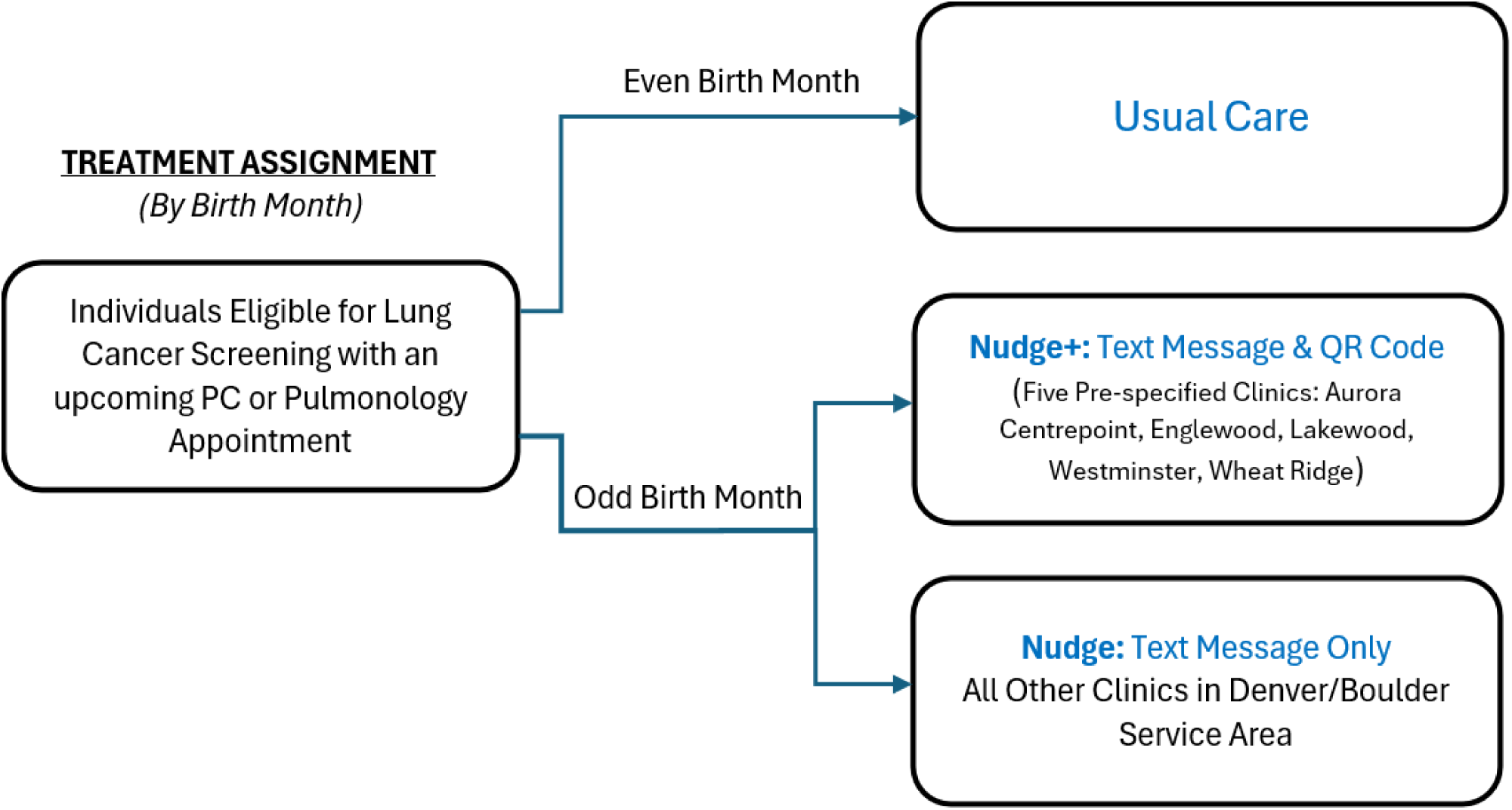
Randomization Process and Intervention Assignment.

### Intervention Methods

A brief LCS informational video (approximately 3 minutes) was developed to describe the benefits, potential harms, and provide information on smoking cessation, and explain the LDCT screening process, including what patients could expect during a CT scan. The video was designed using the EAST framework (Easy, Attractive, Social, Timely) and was narrated by a KPCO pulmonologist to enhance credibility and relevance.^16,17^

Patients in the text-message-only intervention group received a text message the day before an eligible appointment encouraging them to discuss LCS with their provider. Patients in the text-message-plus-QR-code group received the same text message and, during rooming for each eligible visit, were also provided with a QR code linking to the nudge video and encouraged to watch it while waiting for their provider. The text-message intervention was embedded within existing appointment-reminder messages routinely sent to patients, while the QR code workflow was designed to minimize additional burden on clinical staff and providers. Given the pragmatic and feasibility nature of this feasibility study, iterative adaptations were permitted during implementation to improve intervention reach, engagement, and effectiveness.

### Study Outcomes

The study included three primary outcomes: (1) provider-initiated LCS orders, measured both as orders placed on the day of or day following the qualifying visit and as orders placed at any time after the initial visit; (2) completion of a baseline LCS- LDCT; and (3) patient engagement with the intervention, measured by the total number of video views, the number of unique individuals who viewed the video, and the proportion of the video viewed. Because the average wait time for a CT appointment at KPCO was approximately 71 days during the study period, analyses of completed LCS- LDCTs were restricted to patients whose screening order was placed before September 1, 2025, allowing sufficient follow-up time for scan completion.

### Data sources and variables

LCS orders and completed LDCT scans were identified from KPCO radiology data using Current Procedural Terminology (CPT) and Healthcare Common Procedure Coding System (HCPCS) codes G0297, 71271, S8032, 71250, and 71260. Electronic health record (EHR) administrative data were used to identify upcoming primary care and pulmonology appointments, patients with an active or overdue LCS care gap, and patient birth month for study-group assignment. Appointment-level data included visit modality (in-person or virtual), department specialty, and provider years of experience.

Video engagement was measured using QUMU, a secure video-hosting platform used across Kaiser Permanente. Although QUMU does not permit direct linkage of video views to individual patients, a unique device identifier was used as a proxy for distinct viewers. Separate QR codes were created for each of the five participating QR-code clinics, and a distinct video link was used for all text-message outreach, allowing video engagement to be assessed by delivery modality and clinic.

Additional patient-level characteristics were obtained from the KPCO Virtual Data Warehouse (VDW), a standardized, research-ready data resource containing information derived from EHR, administrative, and claims data.^18^ Race and ethnicity were classified into mutually exclusive categories of Asian or Native Hawaiian/Pacific Islander, Black, Hispanic, White, another race, or unknown. Smoking history included smoking status (current or former), pack-years categorized as 20–29, 30–39, or ≥40 pack-years, and, among former smokers, years since quitting categorized as <5, 5–9, 10–14, ≥15 years, or missing. Comorbidity burden was measured using the Deyo adaptation of the Charlson Comorbidity Index based on diagnoses recorded during the year preceding the index date and categorized as 0, 1, 2, or ≥3. Chronic obstructive pulmonary disease (COPD), pneumonia, emphysema, and chronic bronchitis were examined as separate conditions because of their strong clinical correlation to LCS participation. Body mass index (BMI) was categorized as <25, 25–35, or >35 kg/m².

Socioeconomic context was assessed using the Social Vulnerability Index (SVI), a percentile-based measure of community-level social vulnerability, categorized into quintiles, with quintile 1 representing the greatest vulnerability and quintile 5 the least. Prior health care utilization was measured using a binary indicator for any inpatient admission during the year preceding the index date. Health plan type was classified into mutually exclusive categories of HMO, Medicaid, Medicare, or other/unknown.

### Statistical Analysis

Baseline characteristics between intervention and usual care groups were compared using chi-square tests for binary and categorical variables, and t-tests for continuous measures. Intervention engagement was summarized descriptively using the total number of video views; the mean and median proportion of the video viewed; the number and proportion of unique individuals with at least one video view; and the number of views per individual. Differences in video engagement between QR code and text-message delivery modalities were assessed using t-tests for continuous measures and Wilcoxon rank-sum tests for ordinal measures.

LCS order placement was evaluated at two time points: (1) on the day of or day following the qualifying primary care appointment and (2) by the end of the follow-up period. Differences in the proportion of patients receiving an LCS order at each time point were compared between the intervention and usual care groups using chi-square tests. Completion of LCS-LDCT was evaluated using three different denominators: (1) all patients included in the study cohort, (2) patients who received an LCS order, and (3) patients who received an LCS order at least 3 months before the end of the study period to allow sufficient time for LDCT completion. Differences in LCS completion between study groups were assessed using chi-square tests.

Associations between covariates and receipt of an LCS order were evaluated using multivariable logistic regression, adjusting for all covariates listed in Table 1. Adjusted odds ratios (ORs) and 95% confidence intervals (CIs) were estimated to quantify the association between each characteristic and LCS order placement. In the primary analysis, visit modality (in-person vs. virtual) was strongly associated with LCS order placement. To assess the robustness of the intervention effect to visit modality, we conducted a sensitivity analysis repeating the multivariable logistic regression after excluding virtual visits.

**Table 1.** Baseline Characteristics of LCS Eligible Patients by Intervention Assignment.

| Characteristic | Intervention Group | Usual Care | Total | p-value |
| --- | --- | --- | --- | --- |
|  | N (%) | N (%) | N (%) |  |
| Total Patients, N | 549 (50.2) | 544 (49.8) | 1093 (100.0) | N/A |
| Intervention Delivery Mode |  |  |  |  |
| Text Message and QR Code | 316 (57.6) | 0 (0.0) | 316 (28.9) | N/A |
| Text Message Only | 233 (42.4) | 0 (0.0) | 233 (21.3) |  |
| Usual Care (No Intervention) | 0 (0.0) | 544 (100.0) | 544 (49.8) |  |
| Appointment Modality |  |  |  | 0.875 |
| In-Person | 451 (82.1) | 444 (81.6) | 895 (81.9) |  |
| Virtual | 98 (17.9) | 100 (18.4) | 198 (18.1) |  |
| Appointment Department |  |  |  | 0.874 |
| Family Practice | 1 (0.2) | 0 (0.0) | 1 (0.1) |  |
| Internal Medicine | 0 (0.0) | 1 (0.2) | 1 (0.1) |  |
| Primary Care | 547 (99.6) | 542 (99.6) | 1089 (99.6) |  |
| Pulmonary Diseases | 1 (0.2) | 1 (0.2) | 2 (0.2) |  |
| Provider's Years of Experience |  |  |  | 0.539 |
| 0 - 10 years | 86 (15.7) | 73 (13.4) | 159 (14.5) |  |
| 11 - 20 years | 193 (35.2) | 185 (34.0) | 378 (34.6) |  |
| 21 + years | 218 (39.7) | 224 (41.2) | 442 (40.4) |  |
| Missing | 52 (9.5) | 62 (11.4) | 114 (10.4) |  |
| Gender |  |  |  | 0.018 |
| Female | 253 (46.1) | 290 (53.3) | 543 (49.7) |  |
| Male | 296 (53.9) | 254 (46.7) | 550 (50.3) |  |
| Race and Ethnicity |  |  |  | 0.566 |
| Asian/Hawaiian/Pacific Islander | 30 (5.5) | 20 (3.7) | 50 (4.6) |  |
| Black | 30 (5.5) | 22 (4.0) | 52 (4.8) |  |
| Hispanic | 68 (12.4) | 64 (11.8) | 132 (12.1) |  |
| White | 370 (67.4) | 380 (69.9) | 750 (68.6) |  |
| Another Race | 23 (4.2) | 26 (4.8) | 49 (4.5) |  |
| Unknown | 28 (5.1) | 32 (5.9) | 60 (5.5) |  |
| Age Group |  |  |  | 0.816 |
| 50 - 54 | 104 (18.9) | 98 (18.0) | 202 (18.5) |  |
| 55 - 64 | 228 (41.5) | 218 (40.1) | 446 (40.8) |  |
| 65 - 69 | 95 (17.3) | 94 (17.3) | 189 (17.3) |  |
| 70 - 80 | 122 (22.2) | 134 (24.6) | 256 (23.4) |  |
| Smoking Status |  |  |  | 0.506 |
| Current | 269 (49.0) | 278 (51.1) | 547 (50.0) |  |
| Former | 280 (51.0) | 266 (48.9) | 546 (50.0) |  |
| Smoking Pack-Years |  |  |  | 0.557 |
| 20 - 29 pack years | 285 (51.9) | 278 (51.1) | 563 (51.5) |  |
| 30 - 39 pack years | 120 (21.9) | 133 (24.4) | 253 (23.1) |  |
| 40+ pack years | 144 (26.2) | 133 (24.4) | 277 (25.3) |  |
| Time Since Quit<br>(among those who formerly smoked) |  |  |  | 0.296 |
| < 5 years | 79 (14.4) | 93 (17.1) | 172 (31.5) |  |
| 5 - 9 years | 80 (14.6) | 61 (11.2) | 141 (25.8) |  |
| 10 - 14 years | 67 (12.2) | 58 (10.7) | 125 (22.9) |  |
| 15+ Years | 23 (4.2) | 28 (5.1) | 51 (9.3) |  |
| Missing Quit Years | 31 (5.6) | 26 (4.8) | 57 (10.4) |  |
| Previous Non-Lung Cancer Diagnosis |  |  |  | 0.472 |
| No | 515 (93.8) | 504 (92.6) | 1019 (93.2) |  |
| Yes | 34 (6.2) | 40 (7.4) | 74 (6.8) |  |
| Charlson Comorbidity Index |  |  |  | 0.190 |
| 0 | 236 (43.0) | 222 (40.8) | 458 (41.9) |  |
| 1 | 161 (29.3) | 145 (26.7) | 306 (28.0) |  |
| 2 | 64 (11.7) | 87 (16.0) | 151 (13.8) |  |
| 3+ | 88 (16.0) | 90 (16.5) | 178 (16.3) |  |
| Specific Chronic Conditions |  |  |  |  |
| COPD | 132 (24.0) | 130 (23.9) | 262 (24.0) | 1.000 |
| Pneumonia | 19 (3.5) | 18 (3.3) | 37 (3.4) | 1.000 |
| Emphysema | 29 (5.3) | 25 (4.6) | 54 (4.9) | 0.676 |
| Chronic Bronchitis | 1 (0.2) | 1 (0.2) | 2 (0.2) | 1.000 |
| BMI Group |  |  |  | 0.961 |
| < 25 kg/m<sup>2</sup> | 146 (26.6) | 150 (27.6) | 296 (27.1) |  |
| 25-29 kg/m2 | 183 (33.3) | 182 (33.5) | 365 (33.4) |  |
| 30+ kg/m2 | 207 (37.7) | 201 (36.9) | 408 (37.3) |  |
| Missing | 13 (2.4) | 11 (2.0) | 24 (2.2) |  |
| Social Vulnerability Index (Quintile) |  |  |  | 0.572 |
| 1 | 163 (29.7) | 172 (31.6) | 335 (30.6) |  |
| 2 | 118 (21.5) | 93 (17.1) | 211 (19.3) |  |
| 3 | 110 (20.0) | 118 (21.7) | 228 (20.9) |  |
| 4 | 84 (15.3) | 86 (15.8) | 170 (15.6) |  |
| 5 | 72 (13.1) | 74 (13.6) | 146 (13.4) |  |
| Missing | 2 (0.4) | 1 (0.2) | 3 (0.3) |  |
| Inpatient Admission in Year before Outreach |  |  |  | 0.389 |
| No | 506 (92.2) | 493 (90.6) | 999 (91.4) |  |
| Yes | 43 (7.8) | 51 (9.4) | 94 (8.6) |  |
| Payor at time of Appointment |  |  |  | 0.930 |
| HMO | 378 (68.9) | 372 (68.4) | 750 (68.6) |  |
| Medicaid | 43 (7.8) | 40 (7.4) | 83 (7.6) |  |
| Medicare | 115 (20.9) | 121 (22.2) | 236 (21.6) |  |
| Other | 13 (2.4) | 11 (2.0) | 24 (2.2) |  |

## Results

The study cohort included 1,093 individuals, of whom 549 (50.2%) were assigned to the intervention group and 544 (49.8%) to usual care (Figure 2). Baseline characteristics were generally well balanced between groups, including smoking history, appointment modality, and measures of patient health (Table 1). However, a higher proportion of participants in the intervention group were male compared with the usual care group (53.9% vs. 46.7%). Overall, most qualifying appointments were conducted in person (81.9%) and occurred in primary care settings (99.6%). The majority of participants were White (68.6%), aged 50–64 years (69.3%), and had a smoking history of 20–29 pack-years (51.5%).

**Figure 2.**
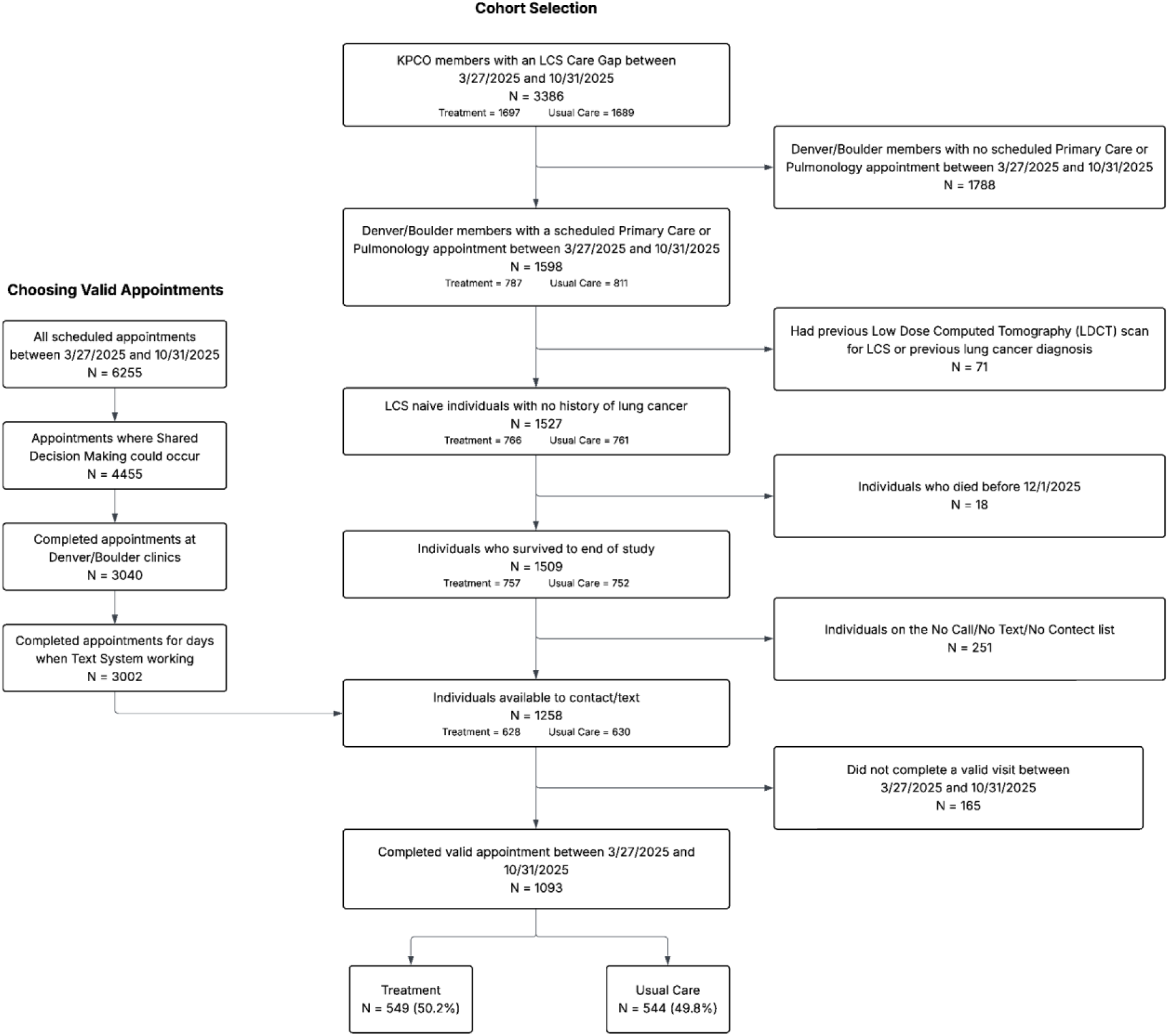
Consort Diagram.

Among the 549 individuals assigned to the intervention group, there were 114 total video views from 93 unique viewers (Table 2). Most video views were accessed through the text-message link rather than the QR code (82.5% vs. 17.5%). Among individuals who viewed the video, most viewed it only once (n=73; 83.8%). Among initiated video views, the mean proportion of the video watched was 78.9%, with a median of 88.0%.

**Table 2.** Video Watches by Delivery Modality.

| Outcome | QR Code Watches | Text Message Watches | Total | p-value |
| --- | --- | --- | --- | --- |
|  | N (%) | N (%) | N (%) |  |
| Unique Patients | 14 (15.1) | 79 (84.9) | 93 | N/A |
| Total Number of Video Watches | 20 (17.5) | 94 (82.5) | 114 | N/A |
| Total Video Watches Per Person |  |  |  | 0.057 |
| 1 | 12 (85.7) | 66 (83.5) | 78 |  |
| 2 | 0 (0.0) | 11 (13.9) | 11 |  |
| 3 | 1 (7.1) | 2 (2.5) | 3 |  |
| 5 | 1 (7.1) | 0 (0.0) | 1 |  |
| Percent of video watched |  |  |  |  |
| Mean, SD | 89.6 (40.7) | 77.0 (38.4) | 78.9 (38.8) | 0.265<sup>a</sup> |
| Median (25th, 75th) | 98.4 (82.8 - 100) | 88 (56.9 - 100) | 88 (56.9 - 100) | 0.414<sup>b</sup> |

<sup>a</sup> t-test<sup>b</sup> wilcoxon

Effects of the nudge intervention on LCS order placement and completion of baseline LCS-LDCT are shown in Table 3. Patients in the intervention group were 38% more likely to receive an LCS order on the day of or day following their appointment compared with those receiving usual care (22.6% vs. 16.4%; p=0.010) and 44% more likely to receive an LCS order at any point during follow-up (32.6% vs. 22.6%; p=0.002). Among all patients in the study cohort, those in the intervention group were 51% more likely to complete a baseline LCS-LDCT (8.6% vs. 5.7%; p=0.078). Among patients who received an LCS order, baseline LCS-LDCT completion was 11% higher in the intervention group (26.3% vs. 23.7%; p=0.691), although this difference did not reach statistical significance. When the analysis was further restricted to patients who received an LCS order at least 3 months before the end of follow-up, completion was 42% higher in the intervention group (35.4% vs. 25.0%; p=0.126); but not statistically significant.

**Table 3.**
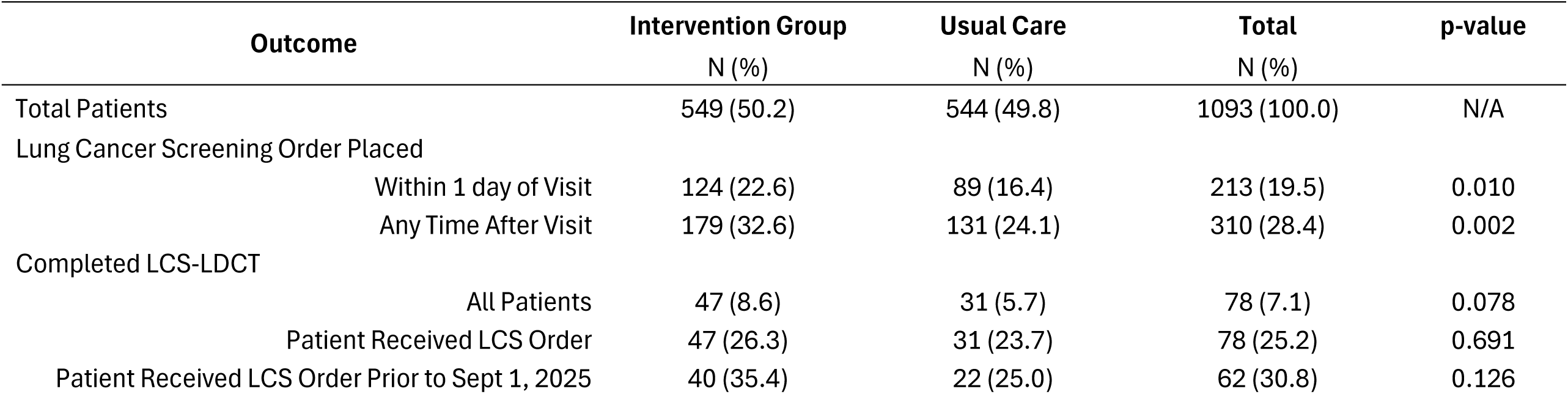
Effect of the LCS Video Nudge Intervention on LCS Orders and Baseline LDCT Completion.

| Outcome | Intervention Group | Usual Care | Total | p-value |
| --- | --- | --- | --- | --- |
|  | N (%) | N (%) | N (%) |  |
| Total Patients | 549 (50.2) | 544 (49.8) | 1093 (100.0) | N/A |
| Lung Cancer Screening Order Placed |  |  |  |  |
| Within 1 day of Visit | 124 (22.6) | 89 (16.4) | 213 (19.5) | 0.010 |
| Any Time After Visit | 179 (32.6) | 131 (24.1) | 310 (28.4) | 0.002 |
| Completed LCS-LDCT |  |  |  |  |
| All Patients | 47 (8.6) | 31 (5.7) | 78 (7.1) | 0.078 |
| Patient Received LCS Order | 47 (26.3) | 31 (23.7) | 78 (25.2) | 0.691 |
| Patient Received LCS Order Prior to Sept 1, 2025 | 40 (35.4) | 22 (25.0) | 62 (30.8) | 0.126 |

Results from the multivariable logistic regression examining factors associated with receipt of an LCS order are shown in Figure 3A. After adjustment for patient and visit characteristics, individuals in the intervention group had 57% higher odds of receiving an LCS order compared with those in the usual care group (OR, 1.57; 95% CI, 1.19–2.07). Visit modality was also strongly associated with LCS ordering; patients receiving virtual care had 70% lower odds of receiving an LCS order compared with those seen in person (OR, 0.30; 95% CI, 0.19–0.48). Sensitivity analysis that restricted to in-person visits is shown in Figure 3B. The intervention effect remained similar, with 51% higher odds of receiving an LCS order compared with usual care (OR, 1.51; 95% CI, 1.13–2.03).

**Figure 3.**
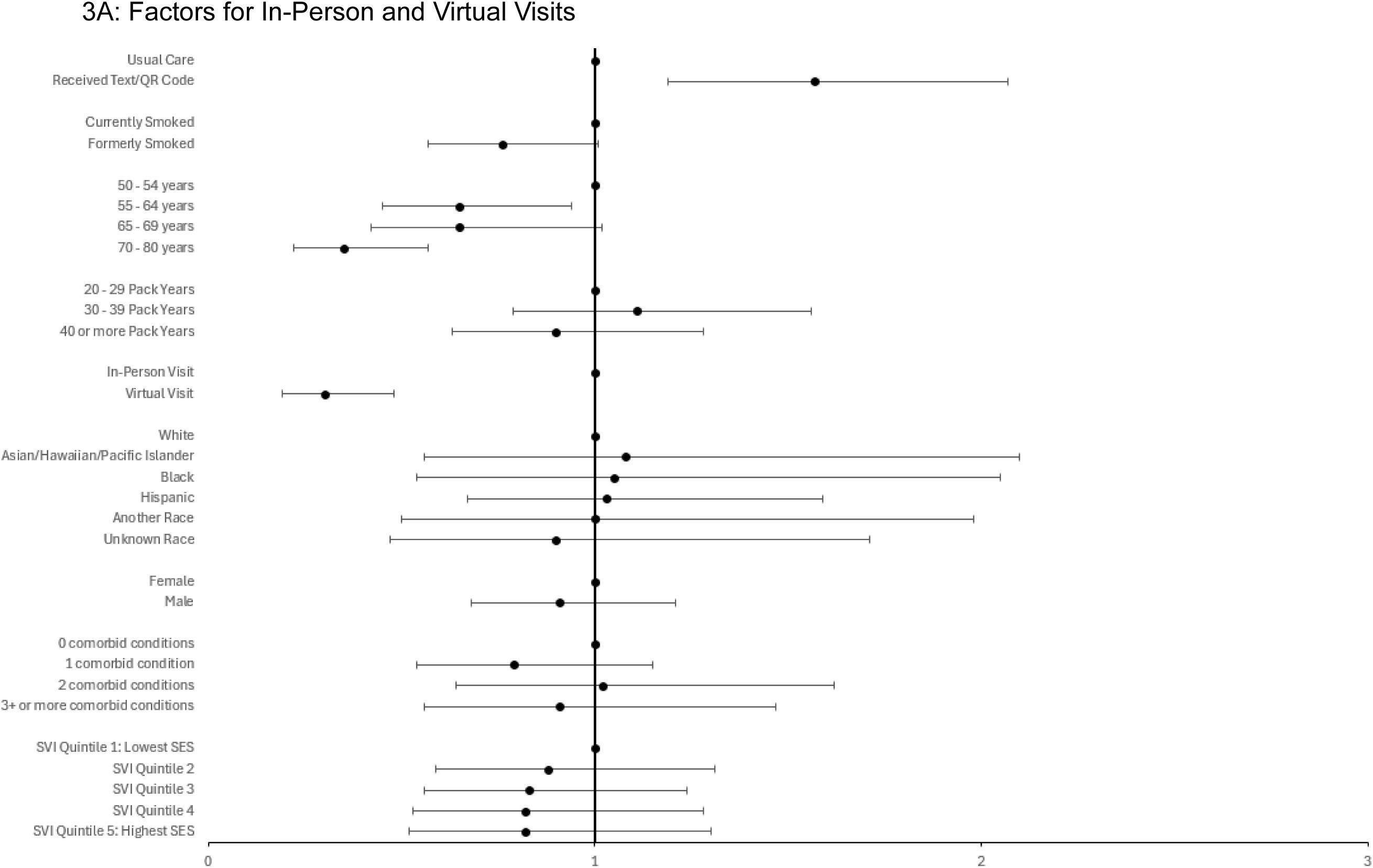
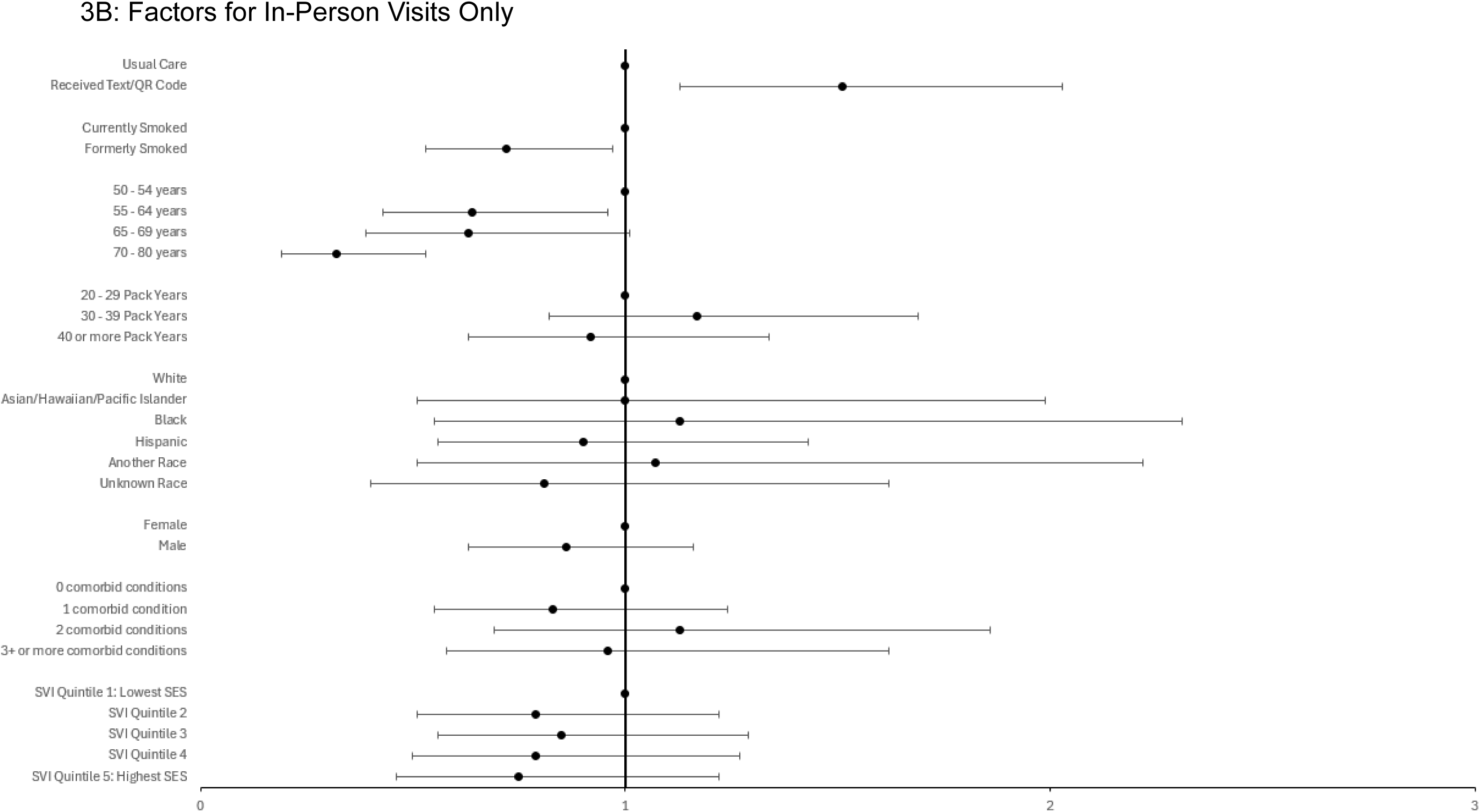
Factors Associated with Receiving a Lung Cancer Screening Order.

## Discussion

In this study, LCS-eligible individuals who received a brief informational video delivered by text message within 24 hours of a scheduled clinical appointment were 44% more likely to receive an LCS order and 51% more likely to complete a baseline LCS-LDCT. Measures of intervention engagement showed that 17% of individuals assigned to the intervention viewed the nudge video and, among viewers, an average of 79% of the video was watched. This level of engagement is particularly notable because the first 80% of the video contained the core behavioral nudge content related to LCS, whereas the final 20% focused primarily on smoking cessation, which we considered a downstream outcome. Taken together, the observed engagement with the intervention and improvements in LCS outcomes support the feasibility and potential effectiveness of this approach. Based on these findings, KPCO incorporated this behavioral nudge intervention into standard clinical care in February 2026.

These findings add to a growing literature of interventions designed to improve LCS participation.^19,20^ Prior studies of video-based decision aids and informational interventions have demonstrated improvements in patient knowledge, informed decision-making, or decisional certainty, but have not consistently translated these improvements into improved LCS uptake among broader populations. For example, a randomized trial of an LCS informational film found improved knowledge and reduced decisional conflict but no difference in screening participation.^13^ In a video-based intervention delivered to a more selected group of LCS-eligible individuals receiving smoking cessation counseling found mixed results. The video was viewed by 14% of intervention participants, but did not significantly increase LDCT utilization in the intention-to-treat analysis, although screening was more common among individuals who actually viewed the video.^14^ More recent digital outreach studies have likewise demonstrated that successfully reaching and engaging patients may be an important determinant of subsequent screening participation.^21^ Our intervention differs from many prior approaches by delivering a brief informational video immediately before a scheduled clinical encounter and embedding the outreach within an existing appointment-reminder workflow. The observed increase in LCS ordering suggests that pairing educational content with a highly actionable opportunity to discuss screening with a provider may help bridge the gap between improving patient knowledge and changing screening behavior.

We also found that intervention delivery modality influenced patient engagement. The vast majority of video views occurred through the text-message link, whereas engagement with the QR code was substantially lower. When designing the intervention, we identified the interval between patient rooming and provider arrival as a potentially valuable opportunity to deliver LCS information at a time when it would remain proximal, salient, and easy to act on during the upcoming encounter.^22^ However, after observing low initial engagement with the QR code, interviews with clinical staff indicated that distributing the QR code fell outside their usual workflow and was not incorporated into routinely monitored performance metrics. As a result, the QR code was often overlooked or ignored during rooming. More fully integrating QR code delivery into established clinical workflows and accountability structures may improve staff uptake and, in turn, increase patient engagement.

### Limitations

This study had several limitations. First, the intervention was implemented over a 9- month period in 2025, which resulted in limited follow-up time for many patients to complete a baseline LCS-LDCT and reduced the precision of estimates for screening completion outcomes. Longer follow-up is also needed to determine whether the observed intervention effects can be sustained over time. Second, although video engagement metrics were available, individual video views could not be directly linked to specific patients. Accordingly, our findings reflect an intention-to-treat effect, and we were unable to estimate the effect of actual video exposure on LCS outcomes. Third, the study was conducted among insured individuals receiving care within an integrated health care delivery system, results may not extend to uninsured populations or patients receiving care in fee-for-service settings. Finally, although baseline characteristics were generally well balanced between study groups, several pragmatic adaptations were made during implementation to improve patient and provider engagement. These adaptations may have influenced intervention reach and effectiveness and should be considered when interpreting the findings.

## Conclusions

In conclusion, our results found that a brief, LCS informational video delivered by text message within 24 hours of a scheduled clinical appointment and embedded within existing appointment-reminder workflows increased provider-initiated LCS ordering and was associated with higher completion of baseline LCS. These findings highlight the potential value of delivering targeted outreach at a highly actionable point in the care pathway, immediately before a clinical encounter, when patients can discuss screening with their provider and act on recommendations in real time. Because the intervention was designed to leverage existing health system infrastructure with minimal additional burden on clinicians, it represents a potentially scalable and efficient strategy for improving LCS participation. Based on findings from this study, KPCO incorporated this behavioral nudge intervention into standard clinical care in February 2026. Future studies should evaluate the durability of these effects over longer follow-up periods and test the intervention across diverse health care systems and patient populations.

## Data Availability

The data underlying this study are not publicly available because they contain protected health information and are subject to Kaiser Permanente Colorado data governance and privacy requirements. Deidentified data may be available from the corresponding author upon reasonable request and subject to institutional review, data use agreements, and applicable regulatory approvals.

